# Apixaban Benefit in Subclinical Atrial Fibrillation Accounting for Competing Risks: A Reanalysis of the ARTESIA Trial

**DOI:** 10.1101/2025.09.19.25335927

**Authors:** Shao-Wei Lo, Darae Ko, Dae Hyun Kim, Long H. Ngo, Daniel E Singer, Sachin J Shah

**Affiliations:** Department of Medicine, Cleveland Clinic Foundation, Cleveland, Ohio, USA; Hinda and Arthur Marcus Institute for Aging Research, Hebrew SeniorLife, Boston, Massachusetts, USA; Department of Medicine, Harvard Medical School, Boston, Massachusetts, USA; Department of Medicine, Beth Israel Deaconess Medical Center, Boston, Massachusetts, USA; Department of Medicine, Massachusetts General Hospital, Boston, Massachusetts, USA

## Abstract

**Background:** It is uncertain if anticoagulants provide a net benefit in subclinical AF (SCAF). The ARTESIA trial showed apixaban reduced the relative hazard of stroke/systemic embolism (SSE) in SCAF, but did not report absolute risk reduction (ARR). The reported Kaplan-Meier analysis did not account for the competing risk of death or 24-hour AF events. We reanalyzed ARTESIA accounting for competing risks to determine the ARR of apixaban vs. aspirin.

**Methods:** Individual time-to-event and time-to-censoring data were extracted from the published Kaplan-Meier curve. ARTESIA classified deaths and 24-hour AF events as censoring events. We probabilistically reclassified them to competing events to estimate the ARR. We used the Aalen-Johansen estimator to estimate the cumulative risk of stroke/systemic embolism at 3.5 years, accounting for competing events. We compared these results to the Kaplan-Meier estimator.

**Result:** After reclassification of deaths and 24-hour AF to competing events, there were 1111 censoring and 852 competing events in the apixaban arm, and 1100 censoring and 816 competing events in the aspirin arm. Compared to the Aalen-Johansen estimator, the Kaplan-Meier estimator overestimated the cumulative SSE risk at 3.5 years with apixaban (2.64% vs. 2.20%, difference in estimates 0.43 percentage points; 95% CI 0.27 to 0.62) and with aspirin (4.58% vs. 3.82%, difference in estimates 0.76 percentage points; 95% CI 0.56 to 1.01). Compared to the Aalen-Johansen estimator, the Kaplan-Meier estimator overestimated the ARR of SSE risk at 3.5 years with apixaban compared to aspirin (1.94% vs. 1.61%, difference in estimates 0.33 percentage points; 95% CI, 0.06 to 0.64).

**Conclusion:** In SCAF, apixaban reduced the 3.5-year risk of SSE by 1.61%, which would be overestimated by 20% if death and >24-hour AF events are treated as censoring rather than competing events.

## Introduction

It is uncertain if anticoagulants provide a net benefit in subclinical atrial fibrillation (SCAF). Apixaban for the Reduction of Thrombo-Embolism in Patients with Device-Detected Subclinical Atrial Fibrillation (ARTESIA) randomized participants with SCAF between 6 minutes and 24 hours and a CHA_2_DS_2_-VASc score ≥2 to either apixaban or aspirin to prevent stroke and systemic embolism (SSE).^1^ When individuals developed >24 hours of AF, they were censored and started on oral anticoagulants. The trial showed apixaban reduced the relative hazard of SSE.

However, ARTESIA did not report absolute risk reduction (ARR), the measure most relevant for clinical decision-making.^2^ The published Kaplan-Meier analysis did not account for the competing risk of death or >24-hour AF events, which were common in ARTESIA, 18% and 24%, respectively. These competing events occurred before the primary outcome and precluded assessing the study’s primary outcome. Failing to account for competing risks overestimates the cumulative risk in each arm and may overestimate the ARR.^3^ We reanalyzed ARTESIA to estimate the ARR of apixaban vs. aspirin, accounting for competing events.

## Methods

ARTESIA’s analytic goal was to measure the causal hazard ratio, treating death and >24-hour AF events as censoring events. To accomplish our goal of measuring the ARR, a different estimand, we used an estimator that treated deaths and >24-hour AF events as competing events—deaths because they precluded the primary outcome, and >24-hour AF events, because they made participants ineligible for the trial.

To conduct the analysis, we extracted time-to-event and censoring data from ARTESIA’s Kaplan–Meier curves using the IPDfromKM package.^4^ In ARTESIA, participants were censored for death, >24-hour AF, dropout, or study end. We reclassified deaths and >24-hour AF from censoring to competing events. Because the trial did not report which censoring events occurred at each time point, we could not directly reclassify deaths and >24-hour AF events. Instead, we probabilistically reclassified these censoring events as competing events. For each arm (a) and year (y), the probability that a censoring event represented a competing event was:

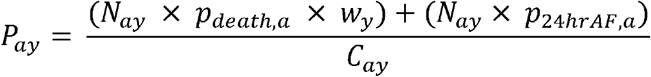

where N=number at risk, p=annual event probabilities, w=Gompertz weight for age-related mortality, and C=total censoring events. We used these probabilities (*P*_*ay*_) to reclassify censoring events as competing events. We repeated this probabilistic reclassification 1,000 times, yielding 1,000 analytic datasets. Results were averaged across these datasets to reflect the uncertainty introduced by probabilistic reclassification.

Using the reclassified data, we plotted the cumulative risk using the Kaplan-Meier estimator, which treated deaths and >24-hour AF events as censoring events, and the Aalen-Johansen estimator,^5^ which treated them as competing events. Then, we used both estimators to calculate the cumulative risk of SSE in each arm and the ARR at 3.5 years, consistent with other secondary analyses of ARTESIA.^6,7^ We calculated the differences in estimates and obtained 95% confidence intervals using 1,000 paired nonparametric bootstrapped replicates of the individual-level data, resampling within each arm and recomputing both the Kaplan–Meier and Aalen–Johansen estimates in each replicate; this ensures the computed confidence intervals accounted for the correlation between estimators.

## Results

The extracted ARTESIA data included 1963 censoring events in the apixaban arm and 1916 in the aspirin arm. After reclassifying deaths and 24-hour AF events as competing events, there were 1111 censoring events and 852 competing events in the apixaban arm, and 1100 censoring events and 816 competing events in the aspirin arm.

Compared to the Aalen-Johansen estimator, the Kaplan-Meier estimator overestimated the cumulative SSE risk at 3.5 years with apixaban (2.64% vs. 2.20%, difference in estimates 0.43 percentage points; 95% CI 0.27 to 0.62) and with aspirin (4.58% vs. 3.82%, difference in estimates 0.76 percentage points; 95% CI 0.56 to 1.01).

Compared to the Aalen-Johansen estimator, the Kaplan-Meier estimator overestimated the ARR of SSE risk at 3.5 years with apixaban compared to aspirin (1.94% vs. 1.61%, difference in estimates 0.33 percentage points; 95% CI, 0.06 to 0.64).

## Conclusion

Competing risk analysis provides a more realistic estimate of patient risks at the point of treatment decision-making.^8^ Using a competing risk analysis, apixaban, compared to aspirin, lowered the absolute SSE risk for people with SCAF by 1.61 percent at 3.5 years. This absolute benefit is overestimated by 20% if death and >24-hour AF events are treated as censoring rather than competing events.

Our study has limitations. Using extracted Kaplan-Meier data necessitated probabilistic identification of competing events, precluded subgroup analysis, and may have introduced approximation errors in event timing. Also, we could not assess how competing risks affect bleeding risk because time-to-bleeding data were unavailable.

Accurate estimation of the ARR is important because, as with clinical AF, the expected absolute reduction in SSE with anticoagulants must be sufficiently large to outweigh their harms. This reanalysis highlights the importance of competing risks methods when measuring the absolute benefit of cardiovascular interventions.

## Data Availability

Original data can be obtained http://doi.org/10.1056/NEJMoa2310234. Analytic data used in this analysis and complete replication of statistical code are available at: https://github.com/MGH-DGIM-SHAH/ARTESIA

## Data sharing

Complete replication of statistical code, including data, is available at: https://github.com/MGH-DGIM-SHAH/ARTESIA

## Ethical review

The Mass General Brigham Institutional Review Board deemed this analysis to be nonhuman subjects research and waived review (#3469).

---

**Figure.**
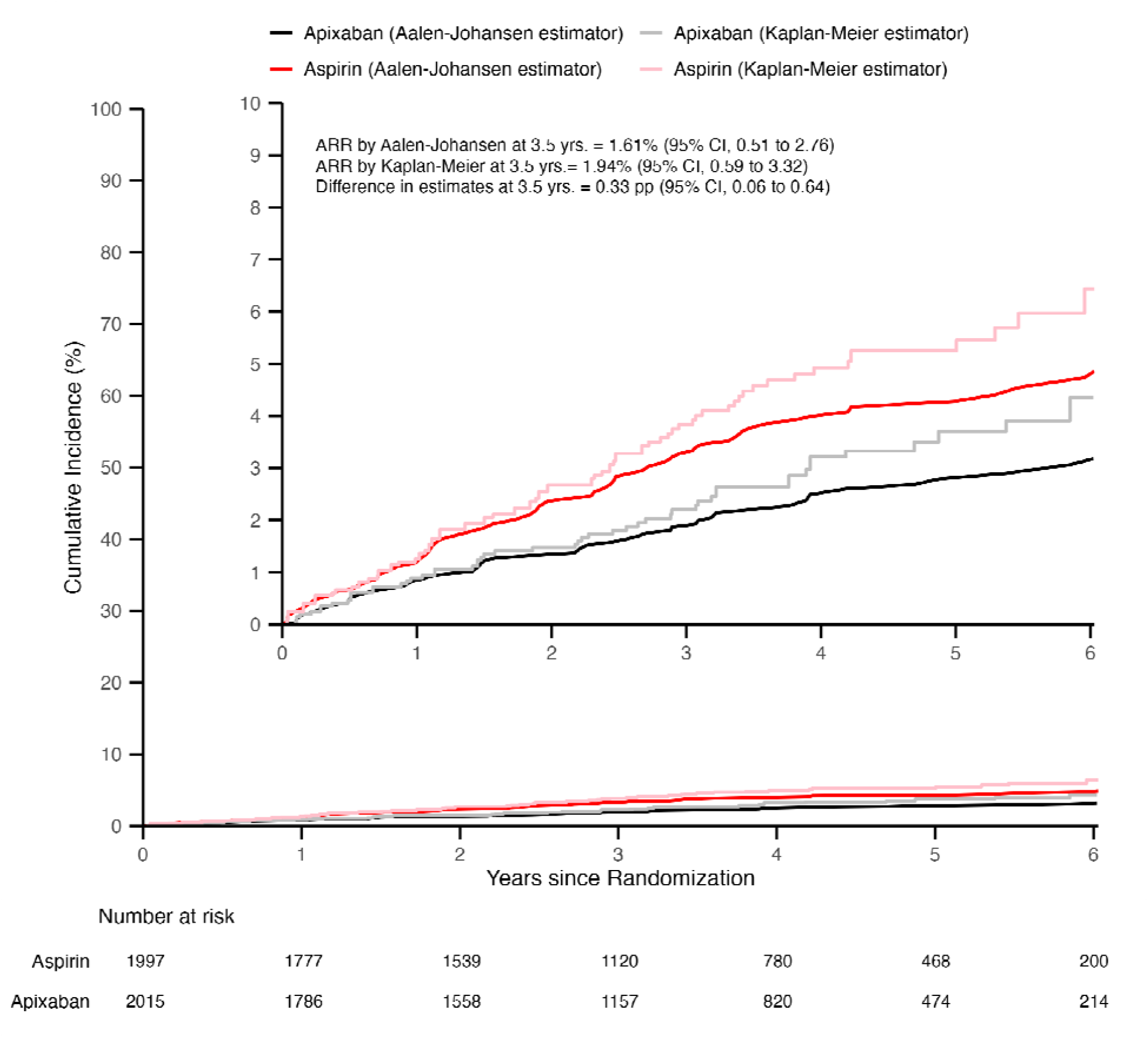
Cumulative incidence of stroke or systemic embolism in the ARTESIA trial, treating death and 24-hour AF events as either censoring events or competing events. ARR – absolute risk reduction, pp – percentage point The figure displays the cumulative incidence of stroke or systemic embolism by study arm, with death and 24-hour AF events treated as censoring events (using a Kaplan-Meier estimator, mirroring the analysis presented in Healey et al., NEJM 2024) and as competing events (using the Aalen-Johansen estimator). The Aalen-Johansen cumulative incidence line is the average across 1000 datasets in which censoring events were probabilistically reclassified as competing events.

